# Edoxaban versus warfarin on stroke risk in patients with atrial fibrillation: a territory-wide cohort study

**DOI:** 10.1101/2021.01.04.21249209

**Authors:** Dicken Kong, Jiandong Zhou, Sharen Lee, Keith Sai Kit Leung, Tong Liu, Abhishek C Sawant, John Corbelli, Abraham KC Wai, Carlin Chang, Qingpeng Zhang, Gary Tse

## Abstract

**Background:** In this territory-wide, observational, propensity score-matched cohort study, we evaluate the development of transient ischaemic attack and ischaemic stroke (TIA/Ischaemic stroke) in patients with AF treated with edoxaban or warfarin.

**Methods:** This was an observational, territory-wide cohort study of patients between January 1^st^, 2016 and December 31^st^, 2019, in Hong Kong. The inclusion were patients with i) atrial fibrillation, and ii) edoxaban or warfarin prescription. 1:2 propensity score matching was performed between edoxaban and warfarin users. Univariate Cox regression identifies significant risk predictors of the primary, secondary and safety outcomes. Hazard ratios (HRs) with corresponding 95% confidence interval [CI] and p values were reported.

**Results:** This cohort included 3464 patients (54.18% males, median baseline age: 72 years old, IQR: 63-80, max: 100 years old), 664 (19.17%) with edoxaban use and 2800 (80.83%) with warfarin use. After a median follow-up of 606 days (IQR: 306-1044, max: 1520 days), 91(incidence rate: 2.62%) developed TIA/ischaemic stroke: 1.51% (10/664) in the edoxaban group and 2.89% (81/2800) in the warfarin group. Edoxaban was associated with a lower risk of TIA or ischemic stroke when compared to warfarin.

**Conclusions:** Edoxaban use was associated with a lower risk of TIA or ischemic stroke after propensity score matching for demographics, comorbidities and medication use.

## Introduction

For decades, vitamin K antagonists (VKAs), such as warfarin, were the only available oral anticoagulants (OACs). While effective in preventing thromboembolism, the VKAs are limited by a narrow therapeutic window, the need for frequent monitoring and extensive drug-drug interactions.(1) Designed to overcome the limitations of the VKAs, the non-vitamin K antagonist oral anticoagulants (NOACs), which have a predictable effect and hence do not require regular monitoring, have revolutionised oral anticoagulation therapy.(2) In the context of ischaemic stroke prevention in patients with atrial fibrillation (AF), all NOACs, namely dabigatran, rivaroxiban, apixaban and edoxaban, have been demonstrated to be at least as effective as warfarin in the landmark NOAC trials. In addition, they demonstrate a significant reduction in intracranial haemorrhage, the most feared bleeding complication of OACs, when compared to warfarin.(3-6) Consequently, there has been a substantial increasing trend in NOAC prescriptions among OAC users with AF, generating an abundance of observational data.(7) Thus far, however, there has been relatively limited observational data for edoxaban due to its late entry among the NOACs. In this territory-wide, observational, propensity score-matched cohort study, we evaluate the development of transient ischaemic attack and ischaemic stroke (TIA/Ischaemic stroke) in patients with AF treated with edoxaban or warfarin. We also consider mortality as a secondary outcome and ischemic heart disease (IHD), hemorrhagic stroke, and gastrointestinal bleeding as safety outcomes.

## Methods

### Study design and population

This was an observational, territory-wide cohort study of patients between January 1^st^, 2016 and December 31^st^, 2019, in Hong Kong. The inclusion were patients with i) atrial fibrillation, and ii) edoxaban or warfarin prescription. The exclusion criteria were patients with i) pulmonary embolus and deep vein thrombosis (PE/DVT), or ii) warfarin use in edoxaban group 90 days before prescription. The remining patients include in total 3464 patients, among them 664 (19.17%) used edoxaban and 2800 (80.83%) used warfarin. The patients were identified from the Clinical Data Analysis and Reporting System (CDARS), a territory-wide database that centralizes patient information from individual local hospitals to establish comprehensive medical data, including clinical characteristics, disease diagnosis, laboratory results, and drug treatment details. The system has been previously used by both our team and other teams in Hong Kong. Patients demographics (gender, baseline age), prior comorbidities (respiratory, renal, endocrine, diabetes mellitus, hypertension, gastrointestinal, congestive Heart failure, intracranial hemorrhage, Peripheral vascular disease, Acute myocardial infarction, Heart failure, baseline IHD, baseline hemorrhagic stroke, baseline gastrointestinal bleeding, and baseline TIA/Ischaemic stroke), medication prescriptions (angiotensin-converting-enzyme inhibitors angiotensin II receptor blockers, calcium channel blockers, beta blockers, diuretics for heart failure, diuretics for hypertension, nitrates, antihypertensive drugs, statins and fibrates, antihyperlipidemic drugs, and anticoagulants), laboratory examinations of potassium, urate, albumin, sodium, urea, protein, creatinine, alkaline phosphatase, aspartate transaminase, alanine transaminase, total bilirubin, HbA1c, APTT and PT were extracted. The list of ICD-9 codes for comorbidity identification are detailed in the **Supplementary Table 1**.

### Outcomes and statistical analysis

The primary outcome was TIA/Ischaemic stroke. Secondary outcome was mortality. Safety outcomes include IHD, hemorrhagic stroke, and gastrointestinal bleeding. Mortality data were obtained from the Hong Kong Death Registry, a population-based official government registry with the registered death records of all Hong Kong citizens linked to CDARS. Descriptive statistics are used to summarize baseline clinical characteristics of patients with TIA/Ischaemic stroke and patients with hemorrhagic presentation. Continuous variables were presented as mean (standard deviation [SD]) or median (interquartile range [IQR]) and categorical variables were presented as count (%). Propensity score matching approach was employed by conducting logistic regression with the treatment group as dependent variable and all confounders as independent variables ^1^, in order to minimize the imbalance arising from confounding between anticoagulants/antiplatelets user cohorts and non-user control groups. Potential confounders included demographics (male gender and baseline age), prior comorbidities (respiratory, renal, endocrine, diabetes mellitus, hypertension, gastrointestinal, congestive heart failure, intracranial hemorrhage, Peripheral vascular disease, Acute myocardial infarction, heart failure, baseline IHD, baseline hemorrhagic stroke, baseline gastrointestinal bleeding, and baseline TIA/Ischaemic stroke), Charlson comorbidity index, and medications (ACEI, Angiotensin II receptor blockers, calcium channel blockers, beta blockers, diuretics for Heart failure, diuretics for hypertension, nitrates, antihypertensive drugs, statins and fibrates, antihyperlipidemic/lipid-lowering drugs, and anticoagulants). We adopt 1:2 nearest-neighbor matching ^2^ to generate control group (warfarin cohort as reference) for the edoxaban cohort. A standardized mean difference (SMD) of less than 0.2 between the treatment groups post-weighting was considered negligible ^3^. Univariate Cox regression identifies significant risk predictors of the primary, secondary and safety outcomes. Hazard ratios (HRs) with corresponding 95% confidence interval [CI] and p values were reported. All significance tests were two-tailed and considered significant if p value<0.05. Statistical analyses were performed using RStudio software (Version: 1.1.456) and Python (Version: 3.6).

## Results

### Basic characteristics

Among the 6957 patients (51.3% males, median baseline age: 67.5 years old, IQR: 59-78, max: 105 years old) with edoxaban or warfarin use from January 1^st^, 2016 to June 30, 2020, in total 2949 non-AF patients, 424 patients with PE/DVT, and 120 patients with warfarin use in edoxaban group 90 days before prescription were excluded (**Figure 1**). The remaining cohort included 3464 patients (54.18% males, median baseline age: 72 years old, IQR: 63-80, max: 100 years old), 664 (19.17%) with edoxaban use and 2800 (80.83%) with warfarin use. After a median follow-up of 606 days (IQR: 306-1044, max: 1520 days), 91(incidence rate: 2.62%) developed TIA/Ischaemic stroke: 1.51% (10/664) in the edoxaban group and 2.89% (81/2800) in the warfarin group. Overall, 663 patients (19.13%) passed away after median follow-up of 614 days (IQR: 313.5-1050, max: 1520 days), 297 patients (8.57%) developed IHD after a median follow-up of 577 days (IQR: 285.5-1018.5, max: 1520 days), 117 patients (3.37%) developed hemorrhagic stroke after median follow-up of 604.5 days (IQR: 306-1043, max: 1520 days), and 112 patients (3.23%) developed gastrointestinal bleeding after median follow-up of 603.5 days (IQR: 306-1043, max: 1520 days). The baseline characteristics between patients with edoxaban use and warfarin use before and after propensity score matching are compared in **Table 1**. Baseline characteristics between patients with TIA/Ischaemic stroke and hemorrhagic stroke before and after propensity score matching are presented in **Supplementary Table 2** and **Supplementary Table 3**, respectively.

**Table 1.** Baseline characteristics of patients with edoxaban/warfarin use before and after propensity score matching (1:2) * for SMD≥0.2

| Characteristics | Before matching |  | SMD | After 1:2 matching |  | SMD |
| --- | --- | --- | --- | --- | --- | --- |
|  | Edoxaban (N=664) | Warfarin (N=2800) |  | Edoxaban (N=612) | Warfarin (N=1380) |  |
|  | Mean(SD);Max;N or<br>Count(%) | Mean(SD);Max;N or<br>Count(%) |  | Mean(SD);Max;N or<br>Count(%) | Mean(SD);Max;N or<br>Count(%) |  |
| **Demographics** |  |  |  |  |  |  |
| Male gender | 339(51.05%) | 1538(54.92%) | 0.08 | 320(52.28%) | 721(52.24%) | 0 |
| Baseline age, year | 75.9(10.6);100.0;n=664 | 69.8(11.8);99.0;n=2800 | 0.55* | 75.7(10.7);100.0;n=612 | 74.4(9.8);99.0;n=1380 | 0.13 |
| **Past comorbidities** |  |  |  |  |  |  |
| Respiratory | 235(35.39%) | 939(33.53%) | 0.04 | 208(33.98%) | 448(32.46%) | 0.03 |
| Renal | 106(15.96%) | 524(18.71%) | 0.07 | 90(14.70%) | 207(15.00%) | 0.01 |
| Endocrine | 8(1.20%) | 25(0.89%) | 0.03 | 5(0.81%) | 19(1.37%) | 0.05 |
| Diabetes mellitus | 36(5.42%) | 247(8.82%) | 0.13 | 31(5.06%) | 75(5.43%) | 0.02 |
| Hypertension | 320(48.19%) | 1004(35.85%) | 0.25* | 292(47.71%) | 598(43.33%) | 0.09 |
| Gastrointestinal | 134(20.18%) | 423(15.10%) | 0.13 | 120(19.60%) | 237(17.17%) | 0.06 |
| Congestive heart failure | 13(1.95%) | 118(4.21%) | 0.13 | 7(1.14%) | 32(2.31%) | 0.09 |
| Hemorrhagic stroke | 22(3.31%) | 107(3.82%) | 0.03 | 20(3.26%) | 45(3.26%) | 0 |
| PVD | 9(1.35%) | 75(2.67%) | 0.09 | 8(1.30%) | 19(1.37%) | 0.01 |
| AMI | 66(9.93%) | 307(10.96%) | 0.03 | 58(9.47%) | 122(8.84%) | 0.02 |
| HF | 217(32.68%) | 1323(47.25%) | 0.30* | 193(31.53%) | 475(34.42%) | 0.06 |
| IHD | 180(27.10%) | 559(19.96%) | 0.17 | 172(28.10%) | 305(22.10%) | 0.14 |
| TIA/Istroke | 41(6.17%) | 163(5.82%) | 0.01 | 41(6.17%) | 72(5.42%) | 0.03 |
| Gastrointestinal bleeding | 26(3.91%) | 49(1.75%) | 0.13 | 21(3.43%) | 49(3.55%) | 0.01 |
| Charlson score | 3.8(1.7);13.0;n=664 | 3.1(1.7);12.0;n=2800 | 0.44* | 3.8(1.6);13.0;n=612 | 3.5(1.4);10.0;n=1380 | 0.17 |
| **Medications** |  |  |  |  |  |  |
| Edoxaban duration, days | 427.6(233.7);2289;n=664 | - | - | 426.9(223.8);1596.0;n=612 | - | - |
| Warfarin duration, days | - | 346.6(131.6);1306;n=2800 | - | - | 345.4(134.9);1306.0;n=1380 | - |
| ACEI | 108(16.26%) | 371(13.25%) | 0.09 | 98(16.01%) | 197(14.27%) | 0.05 |
| ARB | 74(11.14%) | 166(5.92%) | 0.19 | 71(11.60%) | 114(8.26%) | 0.11 |
| Calcium channel blockers | 184(27.71%) | 556(19.85%) | 0.19 | 169(27.61%) | 327(23.69%) | 0.09 |
| Beta blockers | 159(23.94%) | 565(20.17%) | 0.09 | 144(23.52%) | 323(23.40%) | 0 |
| Diuretics for heart failure | 156(23.49%) | 494(17.64%) | 0.15 | 142(23.20%) | 311(22.53%) | 0.02 |
| Diuretics for hypertension | 30(4.51%) | 26(0.92%) | 0.22* | 28(4.57%) | 49(3.55%) | 0.05 |
| Nitrates | 72(10.84%) | 247(8.82%) | 0.07 | 67(10.94%) | 129(9.34%) | 0.05 |
| Antihypertensive drugs | 56(8.43%) | 201(7.17%) | 0.05 | 52(8.49%) | 109(7.89%) | 0.02 |
| Statins and fibrates | 164(24.69%) | 544(19.42%) | 0.13 | 153(25.00%) | 325(23.55%) | 0.03 |
| Antihyperlipidemic | 163(24.54%) | 536(19.14%) | 0.13 | 152(24.83%) | 323(23.40%) | 0.03 |
| Anticoagulants | 275(41.41%) | 1214(43.35%) | 0.04 | 251(41.01%) | 559(40.50%) | 0.01 |
| **Laboratory tests** |  |  |  |  |  |  |
| K/Potassium, mmol/L | 4.2(0.4);5.4;n=608 | 4.1(0.5);6.3;n=1760 | 0.11 | 4.1(0.4);5.4;n=558 | 4.1(0.5);6.3;n=914 | 0.02 |
| Urate, mmol/L | 0.4(0.1);0.9;n=156 | 0.5(0.2);1.1;n=465 | 0.3* | 0.4(0.1);0.9;n=141 | 0.44(0.2);1.1;n=240 | 0.22* |
| Albumin, g/L | 39.1(5.1);50.0;n=595 | 36.5(6.0);51.7;n=2296 | 0.47* | 39.4(4.9);50.0;n=548 | 36.8(6.0);51.7;n=1117 | 0.47* |
| Na/Sodium, mmol/L | 140.3(2.9);147.0;n=608 | 139.4(3.4);154.0;n=1768 | 0.28* | 140.4(2.8);147.0;n=558 | 139.7(3.2);154.0;n=914 | 0.23* |
| Urea, mmol/L | 6.6(2.5);24.8;n=608 | 8.3(5.4);46.0;n=1765 | 0.41* | 6.5(2.4);19.1;n=558 | 8.0(4.7);36.3;n=913 | 0.4* |
| Protein, g/L | 72.5(6.3);92.5;n=590 | 70.8(7.6);99.0;n=2032 | 0.23* | 72.5(6.3);92.0;n=543 | 71.1(7.7);99.0;n=996 | 0.21* |
| Creatinine, umol/L | 94.4(30.5);235.0;n=608 | 132.0(149.2);1536.0;n=1771 | 0.35* | 93.9(29.7);235.0;n=558 | 116.1(108.5);1453.0;n=915 | 0.28* |
| Alkaline phosphatase, U/L | 80.2(36.0);379.0;n=595 | 87.0(40.3);529.0;n=2293 | 0.18 | 79.3(34.3);367.0;n=548 | 85.7(36.0);379.0;n=1115 | 0.18 |
| Aspartate transaminase, U/L | 51.3(201.3);2112.0;n=221 | 38.9(50.7);476.6;n=659 | 0.08 | 43.7(157.2);2112.0;n=195 | 40.9(125.5);2073.2;n=301 | 0.02 |
| Alanine transaminase, U/L | 26.3(46.2);869.2;n=512 | 33.1(55.2);1059.0;n=1857 | 0.13 | 27.0(47.9);869.2;n=472 | 27.1(47.2);1059.0;n=909 | 0 |
| Bilirubin, umol/L | 13.6(8.0);73.0;n=595 | 14.7(11.2);167.2;n=2291 | 0.11 | 13.7(8.0);73.0;n=548 | 14.2(9.9);116.1;n=1114 | 0.05 |
| HbA1c, g/dL | 12.9(2.0);18.9;n=588 | - | - | 13.0(2.0);18.9;n=543 | 12.0(1.9);14.9;n=45 | 0.51* |
| APTT, second | 32.8(7.3);71.9;n=196 | 35.0(9.3);89.4;n=1417 | 0.26* | 33.0(7.5);71.9;n=173 | 34.4(8.5);73.8;n=700 | 0.18 |
| Prothrombin Time, second | 14.9(4.6);43.9;n=176 | 17.0(8.1);108.4;n=1263 | 0.33* | 14.9(4.7);43.9;n=156 | 17.2(8.1);61.0;n=641 | 0.35* |

**Figure 1.**
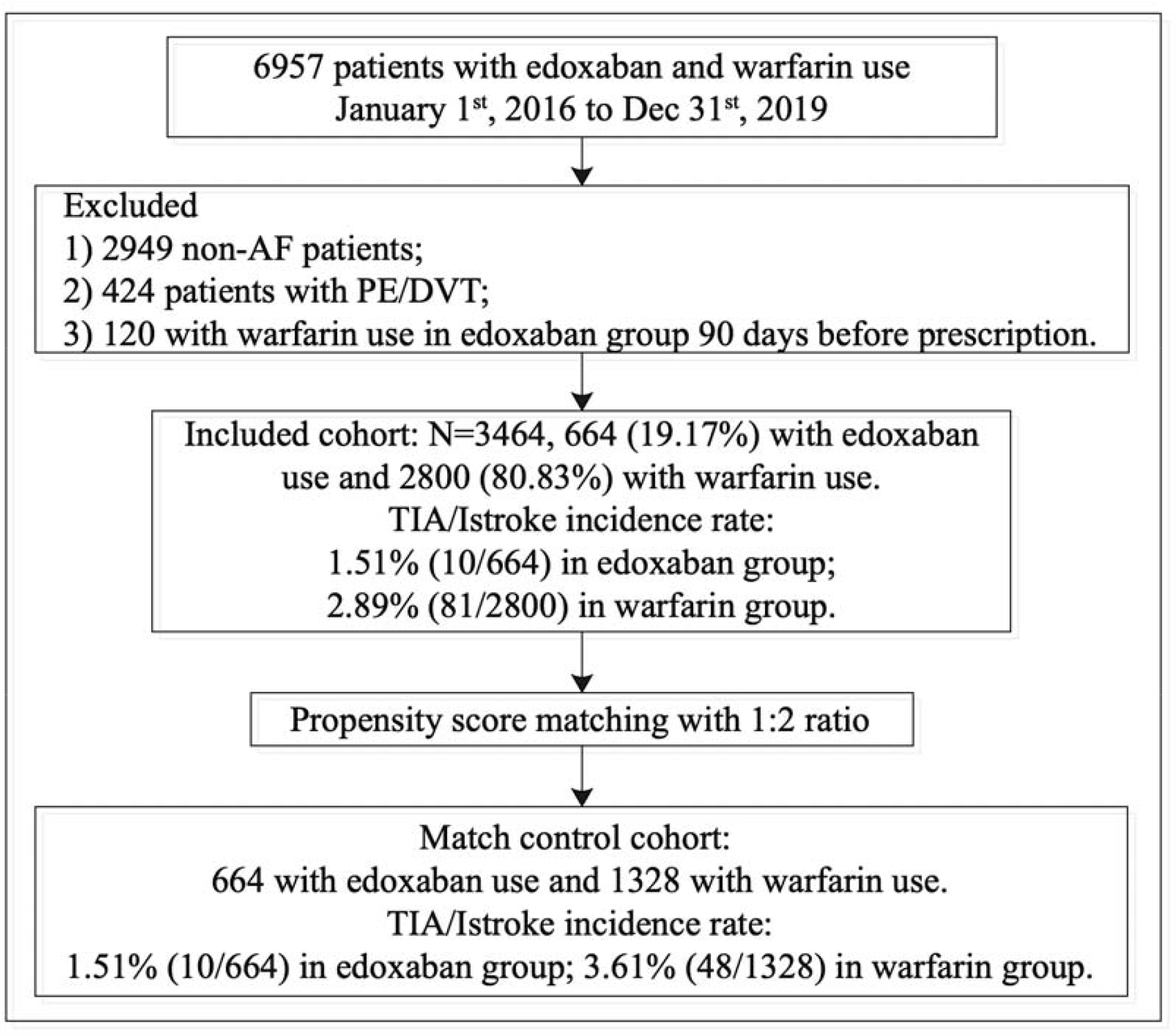
Procedures of data processing.

**Figure 2.**
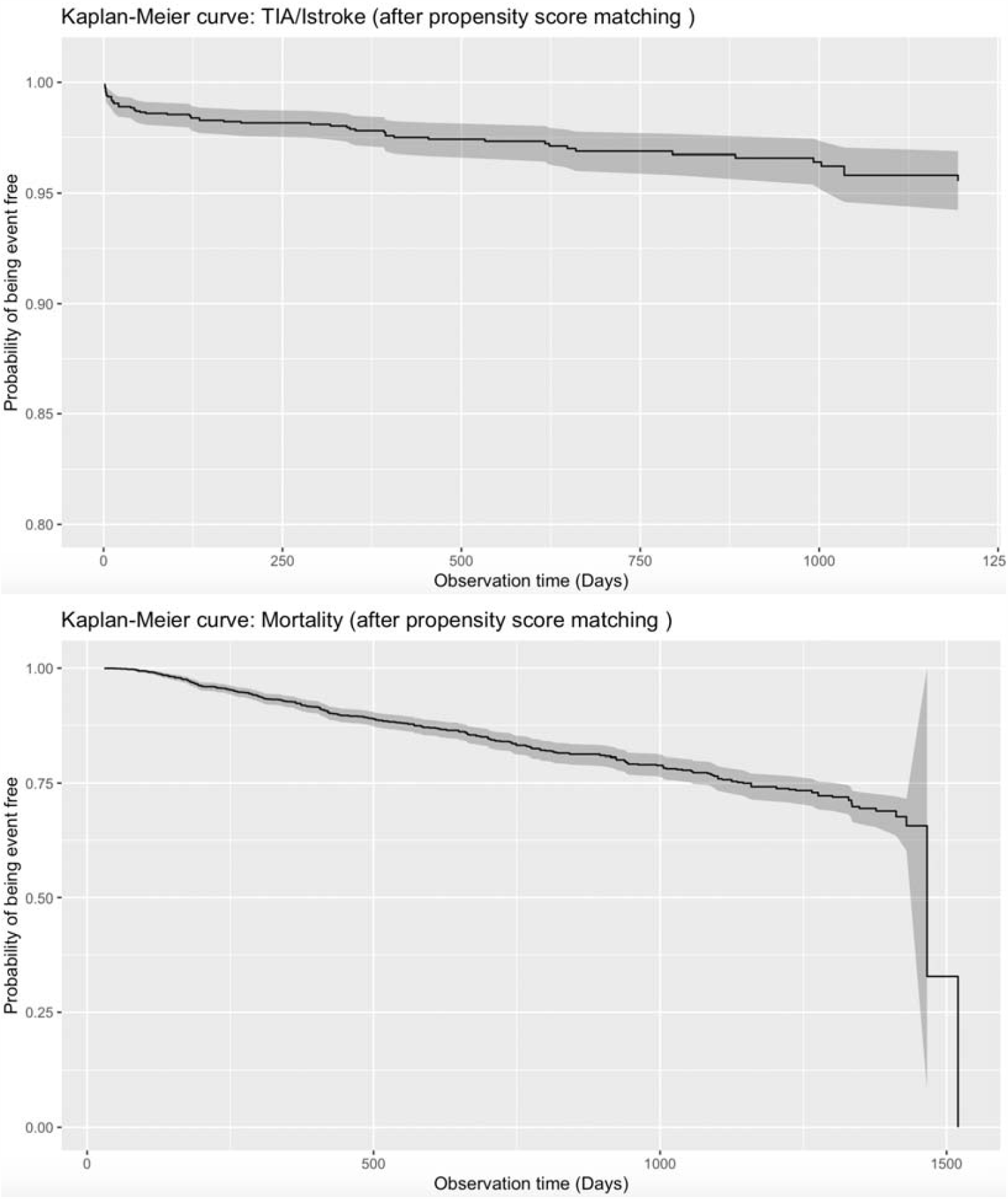
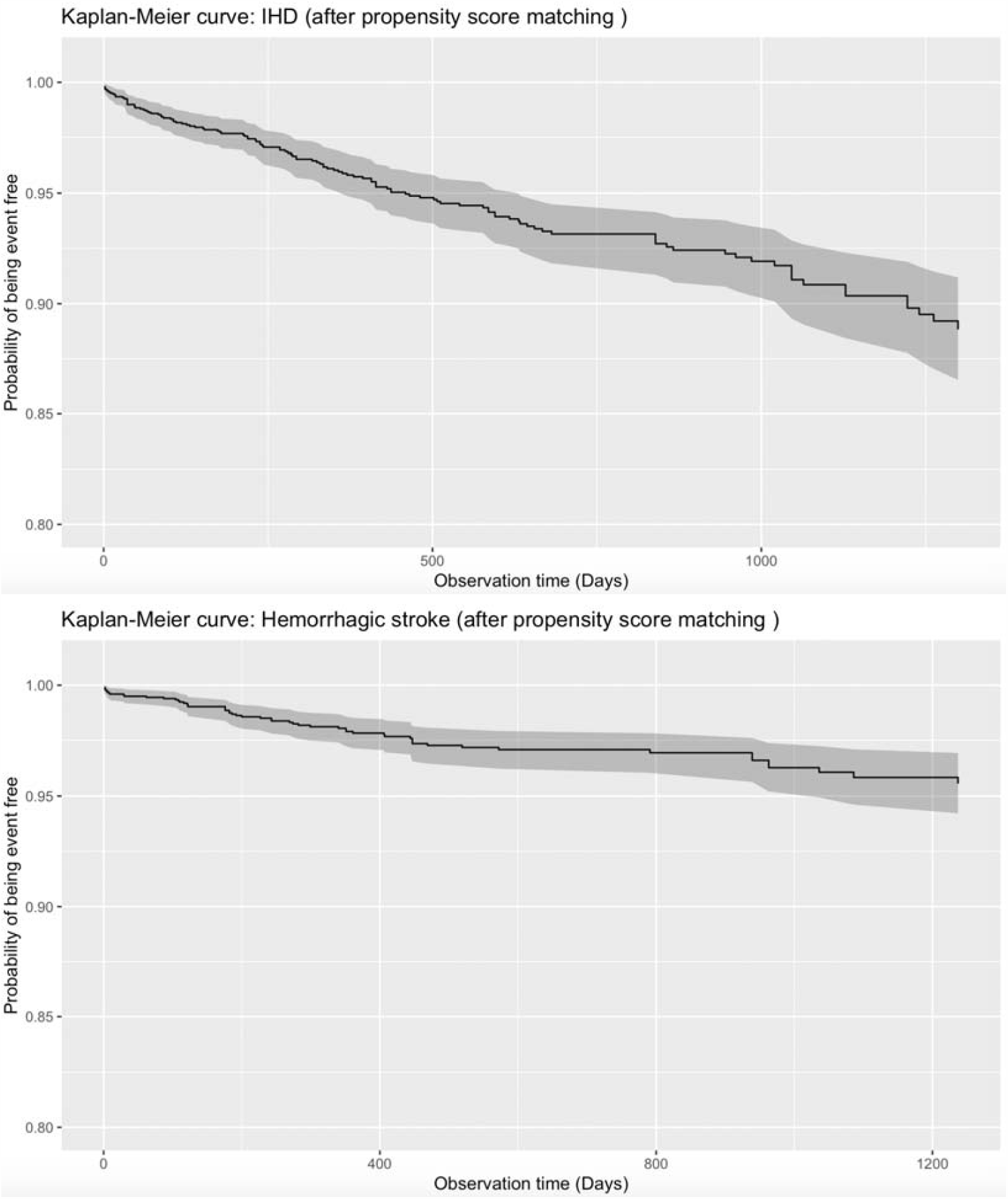
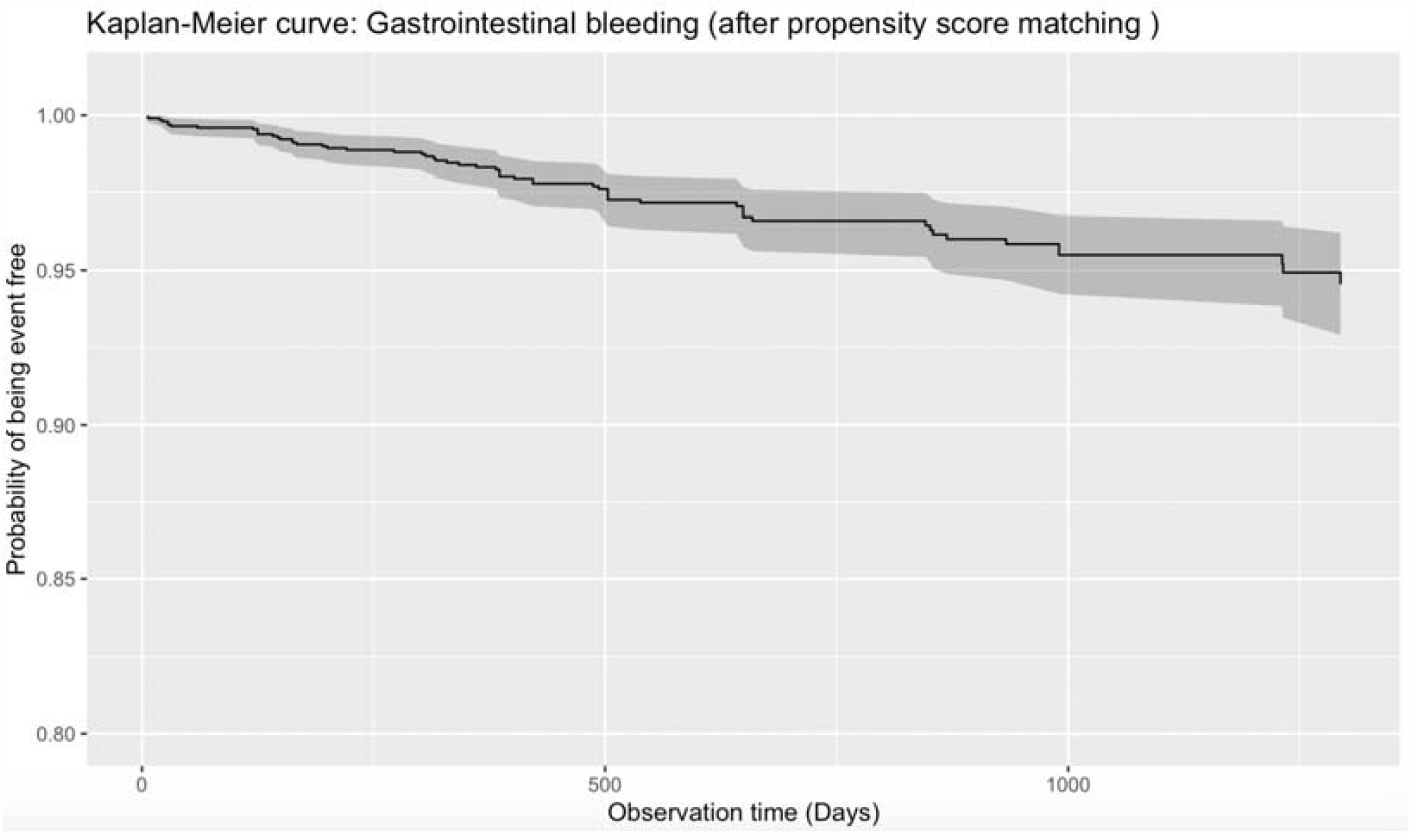
Kaplan-Meier curves of primary, secondary, and safety outcomes.

We now report the statistically significant (i.e. SMD > 0.2) baseline differences between the two groups before matching. For baseline age, the edoxaban users were older (75.93 years v.s. 69.76 years, SMD=0.55) than the warfarin users. Regarding past comorbidities, fewer edoxaban users had heart failure (32.68% v.s. 47.25%, SMD=0.3) while more of them had hypertension (48.19% v.s. 35.85%, SMD=0.25) when compared to warfarin users. Moreover, the edoxaban users had a higher Charlson comorbidity index (3.83 v.s. 3.1, SMD=0.44) than the warfarin users. For prescriptions, a higher proportion of edoxaban users received diuretics for hypertension (4.51% v.s. 0.92%) than did warfarin users, which is hardly surprising considering the fact that more edoxaban users had hypertension. Finally, regarding laboratory test results, the edoxaban users had higher albumin (39.13g/L v.s. 36.54g/L, SMD=0.47), sodium (140.33mmol/L v.s. 139.45mmol/L, SMD=0.28) and protein (72.48g/L v.s. 70.85g/L, SMD=0.23), but had lower urate (0.42mmol/L v.s. 0.46mmol/L, SMD=0.3), urea (6.57mmol/L v.s. 8.29mmol/L, SMD=0.41), creatinine (94.39umol/L v.s. 131.95umol/L, SMD=35), APTT (32.83s v.s. 34.97s, SMD=0.26) and PT (14.86s v.s. 17.02s, SMD=0.33). After 1:2 propensity score matching, statistically significant differences in Charlson comorbidity index, urate, albumin, sodium, urea, protein, creatinine, APTT and PT in remained.

### Signficant risk factors of outcomes

Univariate Cox regression analysis on the propensity score matched cohort identified significantly associated factors of TIA/Istroke (**Table 2**). Edoxaban (warfarin as reference) (HR: 0.10, 95% CI: [0.03, 0.42], P value=0.0015), male gender (HR: 0.57, 95% CI: [0.33, 0.96], P value=0.035), baseline IHD (HR: 0.31, 95% CI: [0.12, 0.77], P value=0.0121), prescription of calcium channel blockers) (HR: 0.43, 95% CI: [0.20, 0.95], P value=0.0363) and higher albumin level (HR: 0.94, 95% CCI: [0.90, 0.98], P value=0.004) were associated with a lower risk of TIA/Istroke. In contrast, prior intracranial hemorrhage (HR: 4.37, 95% CI: [1.58, 12.08], P value=0.004) and baseline hemorrhagic stroke (HR: 3.41, 95% CI: [1.24, 9.42], P value=0.0179) were associated with a higher risk of TIA/Istroke. (delete one of these)

**Table 2.** Univariate predictors of primary TIA/Istroke outcome and secondary mortality outcome after propensity score matching (1:2) * for p≤ 0.05, ** for p ≤ 0.01, *** for p ≤ 0.001

| Characteristics | TIA/Stroke<br>HR [95% CI] | P value | Mortality<br>HR [95% CI] | P value |
| --- | --- | --- | --- | --- |
| Edoxaban (warfarin as reference) | 0.10[0.03, 0.42] | 0.0015 ** | 0.82[0.60, 1.11] | 0.199 |
| Edoxaban duration, days | 0.999[0.996, 1.002] | 0.47 | 0.997[0.996, 0.999] | <0.0001*** |
| Warfarin duration, days | 0.999[0.997, 1.002] | 0.474 | 0.999[0.997, 1.000] | 0.0164* |
| **Demographics** |  |  |  |  |
| Male gender | 0.57[0.33, 0.96] | 0.035* | 1.10[0.89, 1.38] | 0.378 |
| Baseline age, year | 1.02[0.99, 1.05] | 0.17 | 1.09[1.07, 1.11] | <0.0001*** |
| **Past comorbidities** |  |  |  |  |
| Respiratory | 0.59[0.31, 1.12] | 0.106 | 3.05[2.44, 3.80] | <0.0001*** |
| Renal | 0.48[0.17, 1.34] | 0.161 | 2.68[2.08, 3.46] | <0.0001*** |
| Endocrine | 3.67[0.89, 15.09] | 0.0711. | 2.08[0.90, 4.83] | 0.0887. |
| Diabetes Mellitus | 0.34[0.05, 2.47] | 0.288 | 2.52[1.74, 3.63] | <0.0001*** |
| Hypertension | 0.86[0.51, 1.46] | 0.578 | 1.90[1.52, 2.37] | <0.0001*** |
| Gastrointestinal | 0.89[0.44, 1.81] | 0.748 | 1.53[1.18, 1.99] | 0.0014** |
| Congestive heart failure | 0.89[0.12, 6.42] | 0.907 | 3.41[2.14, 5.42] | <0.0001*** |
| Hemorrhagic stroke | 4.37[1.58, 12.08] | 0.004** | 2.69[1.60, 4.51] | 0.0002*** |
| PVD | 1.21[0.17, 8.74] | 0.85 | 1.24[0.55, 2.78] | 0.602 |
| AMI | 0.59[0.18, 1.89] | 0.376 | 2.40[1.78, 3.23] | <0.0001*** |
| HF | 1.26[0.74, 2.15] | 0.389 | 3.15[2.52, 3.94] | <0.0001*** |
| IHD | 0.31[0.12, 0.77] | 0.0121* | 1.33[1.04, 1.70] | 0.0241* |
| Gastrointestinal bleeding | - | - | 2.02[1.30, 3.14] | 0.0019** |
| TIA/Stroke | 1.35[1.03, 1.92] | 0.0011** | 1.94[1.34, 2.81] | 0.0005*** |
| Charlson comorbidity index | 1.07[0.91, 1.27] | 0.414 | 1.46[1.38, 1.54] | <0.0001*** |
| **Medications** |  |  |  |  |
| ACEI | 0.54[0.21, 1.34] | 0.182 | 0.65[0.45, 0.94] | 0.0201* |
| ARB | 0.58[0.18, 1.84] | 0.352 | 0.76[0.48, 1.20] | 0.239 |
| Calcium channel blockers | 0.43[0.20, 0.95] | 0.0363* | 0.71[0.53, 0.95] | 0.0197* |
| Beta blockers | 0.62[0.30, 1.26] | 0.186 | 0.71[0.52, 0.95] | 0.0203* |
| Diuretics for heart failure | 0.56[0.27, 1.19] | 0.131 | 0.55[0.40, 0.76] | 0.0004*** |
| Diuretics for hypertension | - | - | 0.72[0.34, 1.52] | 0.383 |
| Nitrates | 0.73[0.27, 2.03] | 0.551 | 0.55[0.33, 0.92] | 0.0227* |
| Antihypertensive drugs | 0.62[0.19, 1.99] | 0.425 | 1.01[0.67, 1.52] | 0.955 |
| Statins and fibrates | 0.59[0.29, 1.21] | 0.152 | 0.79[0.59, 1.04] | 0.092. |
| Antihyperlipidemic drugs | 0.52[0.25, 1.10] | 0.089. | 0.80[0.60, 1.05] | 0.11 |
| Anticoagulants | 0.62[0.35, 1.10] | 0.0998. | 0.73[0.57, 0.92] | 0.0077** |
| **Laboratory tests** |  |  |  |  |
| K/Potassium, mmol/L | 0.57[0.28, 1.18] | 0.129 | 1.30[1.00, 1.69] | 0.0504. |
| Urate, mmol/L | 2.22[0.08, 61.93] | 0.639 | 10.81[2.86, 40.94] | 0.0005*** |
| Albumin, g/L | 0.94[0.90, 0.98] | 0.0048** | 0.90[0.89, 0.92] | <0.0001*** |
| Na/Sodium, mmol/L | 1.05[0.94, 1.17] | 0.383 | 0.90[0.87, 0.93] | <0.0001*** |
| Urea, mmol/L | 1.04[0.97, 1.11] | 0.304 | 1.10[1.08, 1.12] | <0.0001*** |
| Protein, g/L | 0.98[0.94, 1.01] | 0.229 | 0.96[0.95, 0.97] | <0.0001*** |
| Creatinine, umol/L | 1.00[1.00, 1.00] | 0.751 | 1.003[1.002, 1.004] | <0.0001*** |
| Alkaline phosphatase, U/L | 1.00[1.00, 1.01] | 0.336 | 1.009[1.007, 1.012] | <0.0001*** |
| Aspartate transaminase, U/L | 0.97[0.92, 1.02] | 0.213 | 1.001[1.000, 1.002] | 0.02* |
| Alanine transaminase, U/L | 1.00[0.99, 1.01] | 0.927 | 0.99[0.98, 1.00] | 0.0088** |
| Bilirubin, umol/L | 0.96[0.92, 1.01] | 0.0927. | 1.00[0.98, 1.01] | 0.476 |
| HbA1c, g/dL | 0.96[0.71, 1.30] | 0.784 | 0.75[0.67, 0.84] | <0.0001*** |
| APTT, second | 1.03[0.99, 1.07] | 0.0975. | 1.01[0.99, 1.03] | 0.213 |
| Prothrombin Time/INR, second | 1.02[0.97, 1.06] | 0.502 | 1.01[0.99, 1.03] | 0.225 |

**Table 3.** Univariate predictors of safety outcomes after propensity score matching (1:2) * for p≤ 0.05, ** for p ≤ 0.01, *** for p ≤ 0.001

| Characteristics | IHD<br>HR [95% CI] | P value | Hemorrhagic stroke<br>HR [95% CI] |  | Gastrointestinal bleeding<br>HR [95% CI] | P value |
| --- | --- | --- | --- | --- | --- | --- |
| Edoxaban (warfarin as reference) | 0.35[0.19, 0.66] | 0.0011** | 0.26[0.09, 0.71] | 0.0092** | 1.69[0.95, 3.01] | 0.0766. |
| Edoxaban duration, days | 1.000[0.999, 1.002] | 0.472 | 1.002[1.000, 1.004] | 0.0121* | 1.001[0.999, 1.002] | 0.348 |
| Warfarin duration, days | 1.000[0.999, 1.002] | 0.615 | 1.000[0.997, 1.002] | 0.692 | 0.999[0.997, 1.002] | 0.622 |
| **Demographics** |  |  |  |  |  |  |
| Male gender | 1.27[0.88, 1.82] | 0.196 | 1.50[0.87, 2.59] | 0.141 | 0.76[0.45, 1.27] | 0.295 |
| Baseline age, year | 0.99[0.98, 1.01] | 0.493 | 1.03[1.01, 1.07] | 0.0219* | 1.06[1.03, 1.09] | 0.0003*** |
| **Past comorbidities** |  |  |  |  |  |  |
| Respiratory | 0.83[0.55, 1.24] | 0.36 | 1.47[0.85, 2.54] | 0.167 | 1.32[0.77, 2.27] | 0.317 |
| Renal | 1.39[0.86, 2.25] | 0.181 | 2.64[1.45, 4.79] | 0.0014** | 2.33[1.28, 4.27] | 0.006** |
| Endocrine | 1.94[0.48, 7.85] | 0.354 | 0.00[0.00, Inf] | 0.995 | 2.18[0.30, 15.79] | 0.44 |
| Diabetes mellitus | 2.30[1.27, 4.18] | 0.0062** | 1.60[0.58, 4.43] | 0.366 | 1.58[0.57, 4.36] | 0.379 |
| Hypertension | 1.25[0.88, 1.79] | 0.214 | 0.97[0.57, 1.65] | 0.904 | 1.60[0.95, 2.68] | 0.076. |
| Gastrointestinal | 1.44[0.94, 2.22] | 0.0924. | 0.49[0.19, 1.23] | 0.127 | 1.30[0.69, 2.46] | 0.414 |
| Congestive heart failure | 4.12[2.01, 8.45] | 0.0001*** | 4.21[1.52, 11.64] | 0.0057** | - | - |
| Hemorrhagic stroke | - | - | 21.63[11.79, 39.69] | <0.0001*** | 0.96[0.13, 6.91] | 0.964 |
| PVD | 1.60[0.51, 5.05] | 0.419 | 1.23[0.17, 8.87] | 0.84 | - | - |
| AMI | 3.54[2.32, 5.40] | <0.0001*** | 0.89[0.32, 2.46] | 0.819 | 1.34[0.57, 3.12] | 0.5 |
| HF | 2.26[1.58, 3.22] | <0.0001*** | 1.39[0.81, 2.38] | 0.232 | 1.31[0.77, 2.23] | 0.318 |
| IHD | - | - | 1.04[0.56, 1.93] | 0.912 | 1.68[0.97, 2.91] | 0.0633. |
| Gastrointestinal bleeding | 4.77[2.77, 8.19] | <0.0001*** | - | - | - | - |
| TIA/Istroke | 0.93[0.41, 2.10] | 0.853 | 1.01[0.32, 3.24] | 0.987 | 1.71[0.68, 4.27] | 0.254 |
| Charlson comorbidity index | 1.03[0.91, 1.16] | 0.685 | 1.23[1.05, 1.44] | 0.0102* | 1.21[1.03, 1.42] | 0.0197* |
| **Medications** |  |  |  |  |  |  |
| ACEI | 0.85[0.50, 1.44] | 0.541 | 0.21[0.05, 0.88] | 0.0321* | 1.05[0.52, 2.14] | 0.89 |
| ARB | 0.56[0.25, 1.28] | 0.17 | 1.09[0.43, 2.73] | 0.856 | 0.61[0.19, 1.96] | 0.41 |
| Calcium channel blockers | 0.91[0.59, 1.40] | 0.674 | 0.71[0.36, 1.41] | 0.332 | 0.85[0.45, 1.60] | 0.614 |
| Beta blockers | 1.17[0.78, 1.76] | 0.449 | 0.26[0.10, 0.73] | 0.0101* | 1.10[0.60, 2.00] | 0.768 |
| Diuretics for heart failure | 0.87[0.56, 1.36] | 0.543 | 0.80[0.40, 1.58] | 0.513 | 1.39[0.78, 2.46] | 0.268 |
| Diuretics for hypertension | 0.24[0.03, 1.72] | 0.156 | 1.11[0.27, 4.56] | 0.885 | - | - |
| Nitrates | 1.03[0.55, 1.92] | 0.924 | - | - | 0.57[0.18, 1.81] | 0.337 |
| Antihypertensive drugs | 0.49[0.20, 1.19] | 0.114 | 0.67[0.21, 2.14] | 0.5 | 3.50[1.89, 6.49] | <0.0001*** |
| Statins and fibrates | 1.24[0.83, 1.86] | 0.285 | 0.82[0.42, 1.58] | 0.551 | 1.05[0.58, 1.92] | 0.866 |
| Antihyperlipidemic drugs | 1.26[0.84, 1.88] | 0.258 | 0.83[0.43, 1.60] | 0.575 | 1.07[0.58, 1.95] | 0.836 |
| Anticoagulants | 1.06[0.74, 1.52] | 0.743 | 0.74[0.42, 1.30] | 0.291 | 1.25[0.75, 2.10] | 0.396 |
| **Laboratory tests** |  |  |  |  |  |  |
| K/Potassium, mmol/L | 1.46[0.95, 2.23] | 0.0821. | 0.72[0.35, 1.48] | 0.367 | 0.54[0.28, 1.02] | 0.0581. |
| Urate, mmol/L | 3.52[0.29, 42.22] | 0.32 | 0.64[0.02, 18.47] | 0.792 | 2.64[0.10, 68.50] | 0.56 |
| Albumin, g/L | 0.98[0.95, 1.01] | 0.179 | 0.95[0.91, 0.99] | 0.0175* | 0.93[0.89, 0.97] | 0.0007*** |
| Na/Sodium, mmol/L | 1.03[0.96, 1.10] | 0.464 | 0.91[0.82, 1.00] | 0.0466* | 1.08[0.98, 1.19] | 0.135 |
| Urea, mmol/L | 1.03[0.99, 1.08] | 0.139 | 1.08[1.02, 1.14] | 0.0076** | 1.05[0.99, 1.12] | 0.118 |
| Protein, g/L | 0.97[0.94, 0.99] | 0.007** | 0.97[0.94, 1.01] | 0.186 | 0.98[0.95, 1.02] | 0.374 |
| Creatinine, umol/L | 1.002[1.001, 1.003] | 0.0062** | 1.002[1.000, 1.004] | 0.0487* | 1.00[1.00, 1.00] | 0.676 |
| Alkaline phosphatase, U/L | 0.998[0.991, 1.004] | 0.503 | 1.005[0.999, 1.011] | 0.128 | 1.01[1.00, 1.01] | 0.0183* |
| Aspartate transaminase, U/L | 0.997[0.985, 1.010] | 0.674 | 0.996[0.977, 1.016] | 0.691 | 0.98[0.95, 1.02] | 0.289 |
| Alanine transaminase, U/L | 0.995[0.984, 1.007] | 0.443 | 1.001[0.995, 1.006] | 0.792 | 0.98[0.95, 1.00] | 0.0662. |
| Bilirubin, umol/L | 1.000[0.979, 1.021] | 0.993 | 0.99[0.96, 1.03] | 0.581 | 0.99[0.95, 1.02] | 0.395 |
| HbA1c, g/dL | 0.89[0.75, 1.06] | 0.206 | 0.86[0.61, 1.23] | 0.411 | 0.77[0.62, 0.95] | 0.0169* |
| APTT, second | 0.99[0.96, 1.02] | 0.562 | 0.98[0.93, 1.03] | 0.328 | 1.07[1.04, 1.10] | <0.0001*** |
| Prothrombin Time/INR, second | 0.97[0.92, 1.02] | 0.19 | 0.95[0.87, 1.03] | 0.221 | 1.05[1.02, 1.08] | 0.0003*** |

The risk factors of mortality (**Table 2**) include (1) older baseline age (HR: 1.09, 95% CI: [1.07, 1.11], P value<0.0001); (2) comorbidities of respiratory diseases, renal diseases, diabetes mellitus, hypertension, gastrointestinal diseases, congestive heart failure, intracranial hemorrhage, acute myocardial infarction, heart failure, baseline IHD, baseline hemorrhagic stroke, baseline gastrointestinal bleeding, baseline TIA/Ischaemic stroke and a larger Charlson comorbidity index (HR>1, P value<0.05); (3) high levels of urate, urea, creatinine,, alkaline phosphatase, and aspartate transaminase (HR>1, P value<0.05), and lower levels of albumin, sodium, protein, alanine transaminase and HbA1c (HR<1, P value<0.05). Prescriptions of ACEI, calcium channel blockers, beta blockers, diuretics for heart failure, nitrates, and anticoagulants were protective against mortality (HR<1, P value<0.05). In addition, longer prescription duration of both edoxaban (HR: 0.997, 95% IC: [0.996, 0.999], P value<0.0001) and warfarin (HR: 0.999, 95% CI: [0.997, 1.000], P value=0.0164) were associated with a lower mortality risk. Associations of prescription duration of edoxaban and warfarin with mortality risk are presented in **Figure 4**.

**Figure 3.**
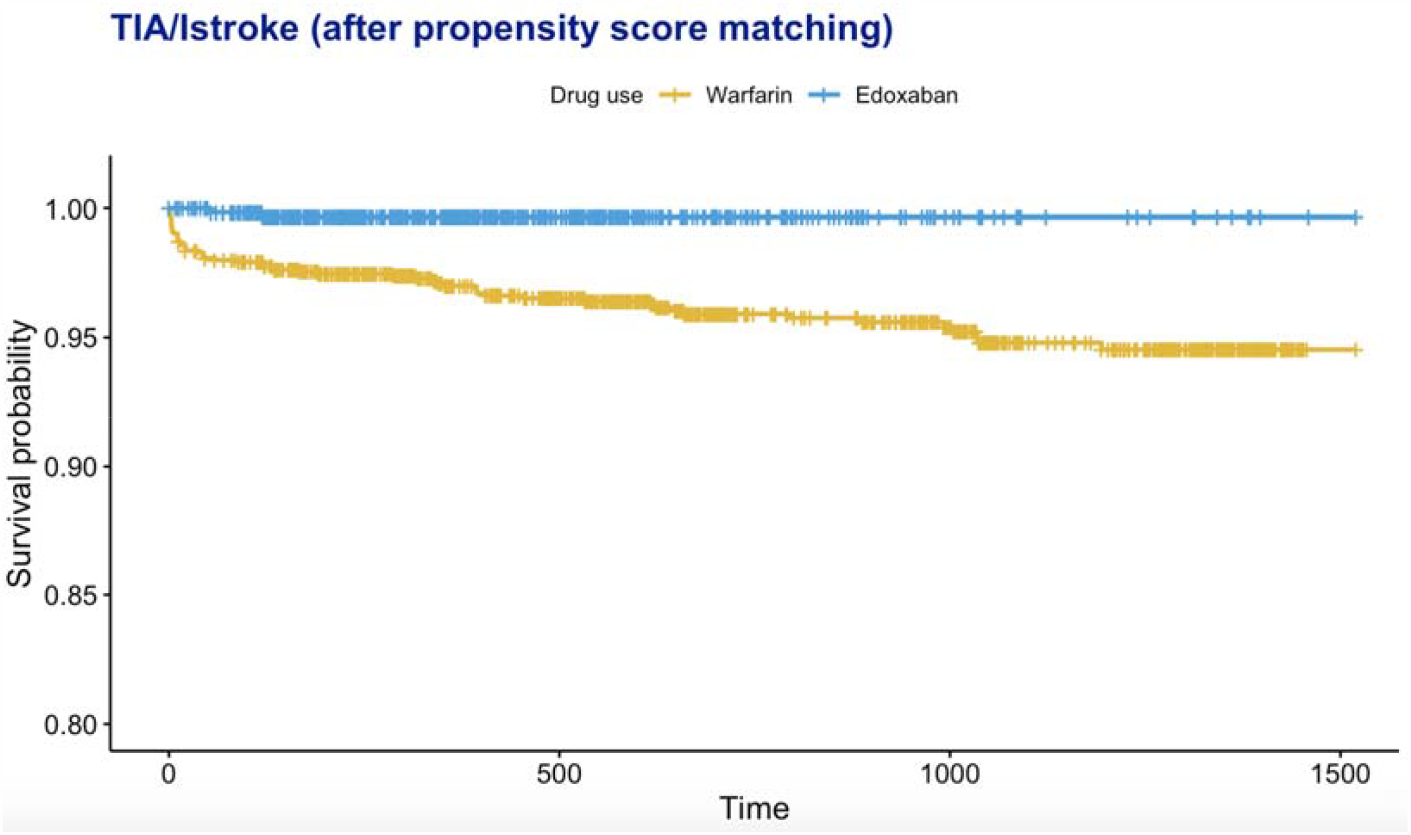
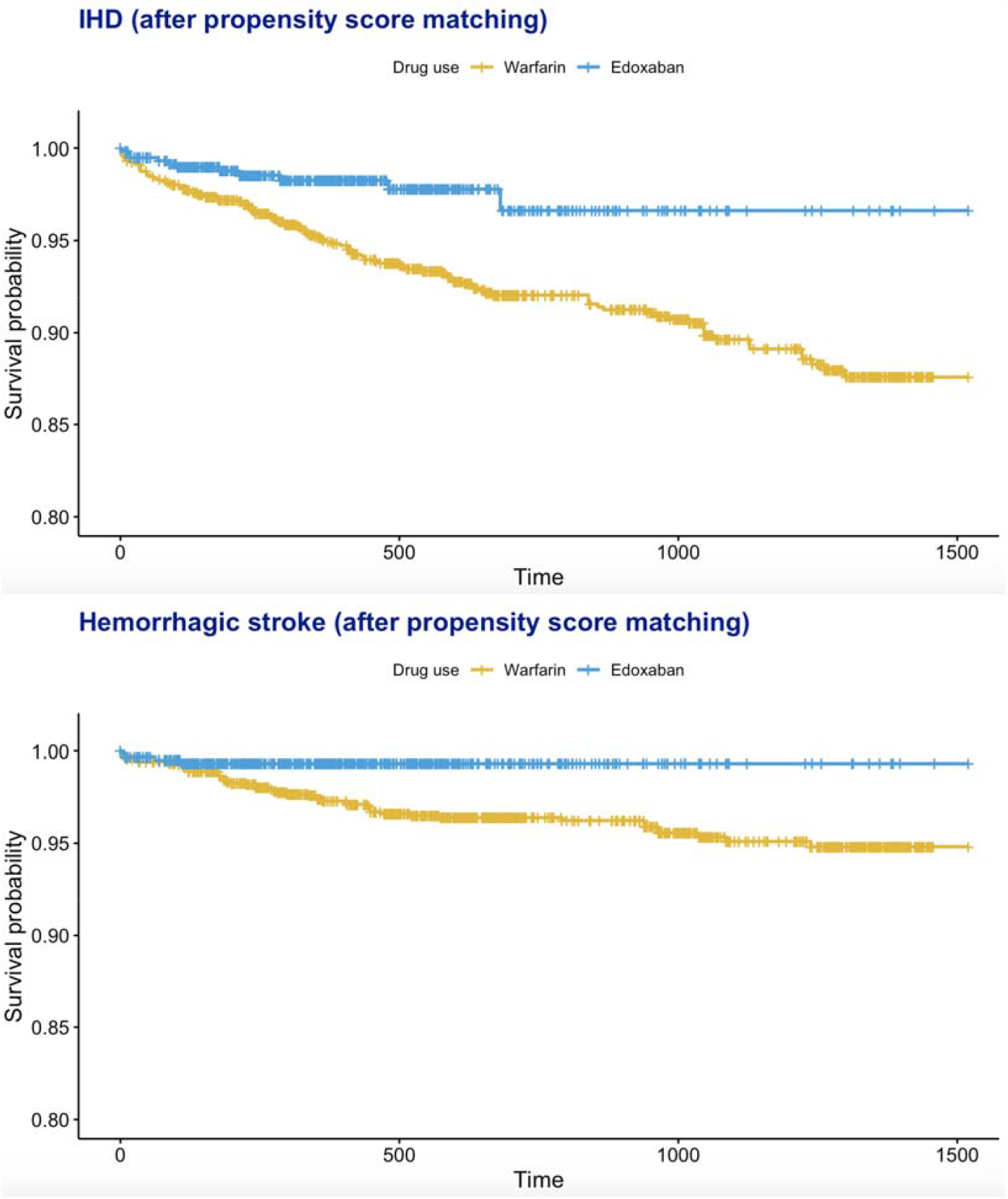
Kaplan-Meier curves of TIA/Istroke, IHD, and hemorrhagic stroke stratified by prescriptions of warfarin and edoxaban drugs.

**Figure 4.**
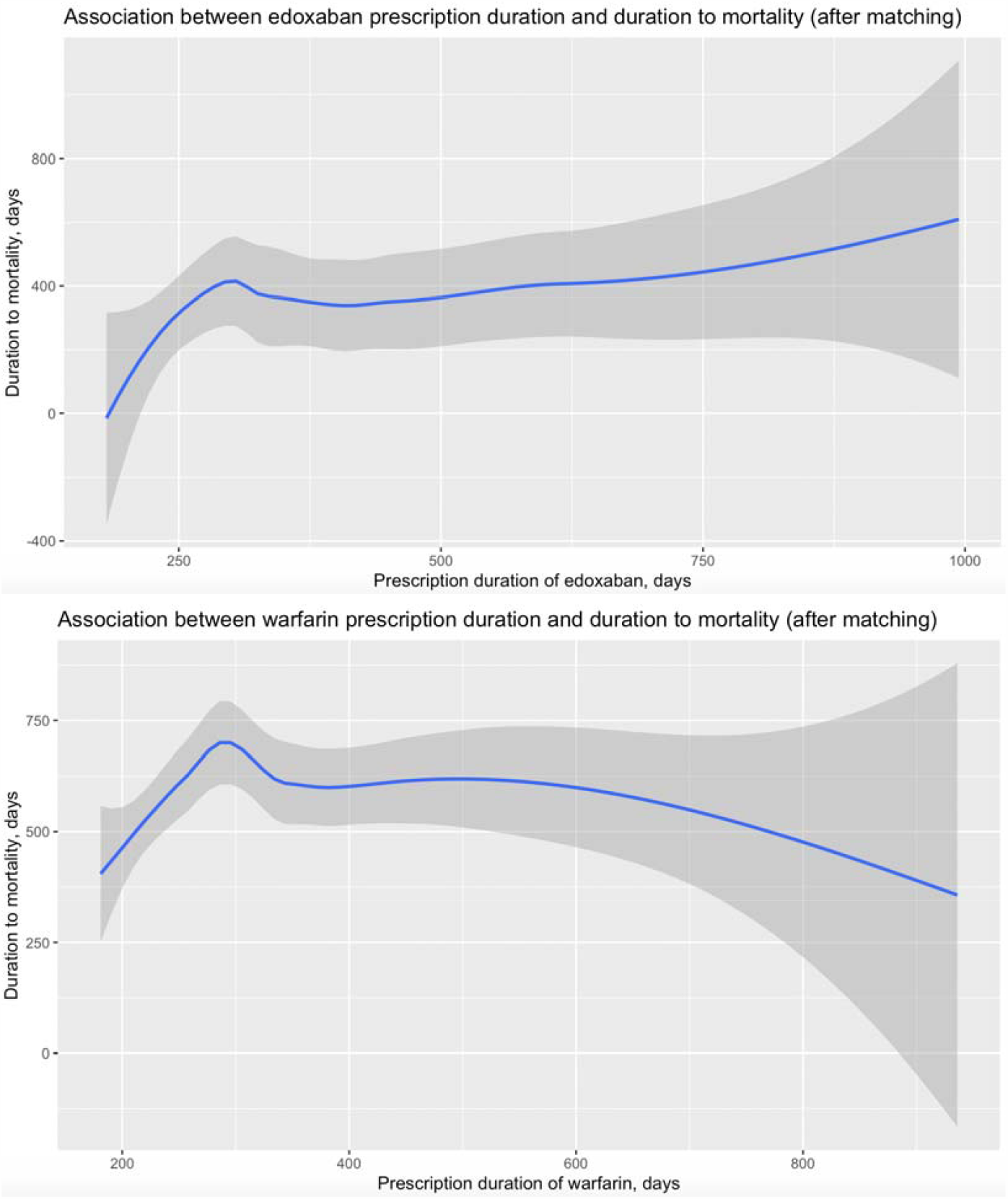
Association of edoxaban/warfarin prescription duration with duration to mortality after propensity score matching (1:2).

With regarding to the safety outcomes, prior comorbidities of diabetes mellitus, congestive Heart failure, Acute myocardial infarction, Heart failure, lower protein level, and higher creatinine level demonstrates significant predictive power for IHD presentation. Edoxaban (warfarin as reference) was associated with lower risk of IHD (HR: 0.35, 95% CI: [0.19, 0.66], P value=0.0011). Older baseline age, prior renal diseases, prior congestive Heart failure, prior intracranial hemorrhage, larger Charlson comorbidity index, lower albumin level, lower sodium level, larger urea amount, and higher creatinine level are significant predictive for hemorrhagic stroke presentation. Medication use of ACEI and beta blockers were protective against hemorrhagic stroke (HR<1, P value<0.05). Prescription of edoxaban (warfarin as reference) (HR: 0.26, 95% CI: [0.09, 0.71], P value=0.0092) with longer duration (HR: 1.002, 95% CI: [1.000, 1.004], P value=0.0121) were associated with lower development risk of hemorrhagic stroke. Finally, older baseline age, prior renal diseases, larger Charlson comorbidity index, lower albumin level, higher alkaline phosphatase level, lower Glycated hemoglobin (HbA1c) level, longer APTT, and longer PT were significant predictive for gastrointestinal bleeding risk. Prescription of antihypertensive drugs showed adverse effects on development of gastrointestinal bleeding.

## Discussion

The main finding of this territory-wide observational study was that edoxaban was associated with a lower risk of TIA or ischemic stroke when compared to warfarin. In contrast, the pivotal ENGAGE AF-TIMI 48 trial found that high-dose edoxaban and warfarin carried a nondifferent risk of ischaemic stroke while low-dose edoxaban gave rise to a higher risk of ischemic stroke than did warfarin.(3) While one may suspect the discrepancy here is attributable to racial differences, as the majority of the ENGAGE AF-TIMI 48 population were non-Asians, an ENGAGE AF-TIMI 48 sub-analysis on East Asian patients showed that the same results remained true among the East Asian patients of the trial.(8) The underlying reason is thus likely a result of the fundamental differences between clinical trials and real life. Importantly, in the ENGAGE AF-TIMI 48 trial, the international normalised ratio (INR) was well-managed among the warfarin group: the median time in the therapeutic range (TTR) was 68.4% of the treatment period.(3) In real life, however, Asians are less likely to achieve optimal INR control due to an increased bleeding tendency during warfarin treatment.(9) Our finding that edoxaban is associated with a lower risk of TIA/stroke is consistent with several observational studies performed in Asia.(10,11,12) However, although edoxaban was associated with a reduced all-cause death in a Korean observational study,(12) we found a non-statistically significant reduction in mortality in our study.

Regarding the safety outcomes, we found that edoxaban was associated with reduced risks of both haemorrhagic stroke and IHD. Although some observational studies in Asia have similarly reported that the NOACs were associated with a lower risk of intracranial haemorrhage when compared to warfarin,(10, 12) the risks of IHD associated with the use of different OACs are not frequently discussed. An additional noteworthy observation in our study was that the risk of haemorrahgic stroke increased with the duration of edoxaban prescription. Whether the safety profile of edoxaban, and indeed of the NOACs in general, depends on treatment duration is a clinically relevant question that requires further studies to explore. For gastrointestinal bleeding risk, we found a non-statistically significant increase associated with edoxaban. This finding is inconsistent with some Korean observational studies, which found that, edoxaban carried smaller risks of gastrointestinal bleeding and hospitalisation for gastrointestinal bleeding than did warfarin(10,11). Further observational studies comparing gastrointestinal bleeding risks associated with warfarin and edoxaban are necessary to improve our understanding on this clinically relevant issue.

### Limitations

An important and well-known limitation of the use of propensity scores is that it is unlikely all possible confounders are accounted for, (13) as we are only able to include data that are available on CDARS in our analysis. As we compare edoxaban and warfarin in this study, we would like the patients to be perfectly compliant to their prescriptions and, for those taking warfarin, to be well managed with regular TTR measurements. Unfortunately, such enforcements are not possible in a retrospective study like ours. Importantly, if the quality of warfarin therapy were low, it may have been responsible for the better performance of edoxaban in reducing TIA/Ischaemic stroke in our findings. In addition, there may be some degree of miscoding and under-reporting of events on CDARS. Finally, the majority of patients on CDARS are of Chinese ethnicity; thus, our results may not be generalizable to other populations.

## Data Availability

Data available upon request.

