## Supplementary Appendix for "Edoxaban versus warfarin on stroke risk in patients with atrial fibrillation: a territory-wide cohort study"

**Supplementary Table 1.** Codes for commodities.

| **Comorbidity** | **Codes** |
| --- | --- |
| Respiratory | 786.09, 518.81, 780.53, 137, E912, 465.9, 518.81, 518.81, 79.6, 518.81, 519.8, 780.59, 799.1, 780.57, 518.82, 480.1, 786.3, 519.8, 997.3, 165.9, 519.9, 648.91, 162.9, 162.3, 197, 162.5, 162.4, 486, 518.89, 496, 162.8, 415.1, V10.11, 518, 162.9, 11.96, 482.1, 507, 513, 11.9, 511.8, 511.1, 11.94, 516.8, 793.1, 482.4, 507, 515, 197, 11.93, 482, 482.83, 518.4, 482.3, 482.2, 415.1, 502, 518.89, 235.7, 793.1, 934.8, 516.9, 136.3, 38.49, 506, 112.4, 487, 481, 117.9, 38.2, 518, 11.95, 79.89, 518.1, 480.9, 505, 516.8, 495.9, 518.3, 11.23, 416.8, 513, 397.1, 117.3, 483, 508, 998.81, 416, 514, 861.21, 502, 934.8, 480.8, 648.93, 11.2, 492.8, 484.6, 78.5, 484.1, 516.3, 415.1, 416.9, 415, 416.9, 429.89, 415, 747.49, 745, 417, 770.7, 427.5, 416.9, 416, 416.8, 746.02, 573.8, 642.9, 416, 747.3, 747.3, 770.3, 779.8, 515, 424.3, 416, 417.8, 747.3, 747.3, 745.4, 518.81, 786.09, V12.6, 478, 748.5, 162.9, 996.84, 748.5, 748.6, V42.1, 748.5, 11.05, 162, 518, 747.42, 518.89, 748.5, 517.2 |
| Renal | 198.7, 189, 189, 585.9, V56.0, 189.1, 584.9, 189.1, 593.9, 189, 189, 189.8, 239.5, 189, 583.81, 593.9, 591, V10.52, 250.4, 591, 255.4, 590.8, 586, 585.1, 591, 189, 591, 239.7, 788, 996.39, 189.1, 588.9, 592, 996.39, 250.4, 593.2, 255.4, 572.4, 194, 255, 453.3, 198, 585.9, 996.39, 996.39, 590.1, 591, 591, 223, 591, 584.9, 585.9, 250.41, 996.39, 581.9, 227, 593.5, 583.89, 593.89, 404.93, 255.5, 788.9, 250.43, 227, 405.92, 592, 753.12, 996.39, E879.1, V42.0, 592, 580.89, 403.9, 593.9, V59.4, 585.9, 580.9, 250.41, 996.39, 585.9, 794.4, 584.8, 584.5, 255.9, 441.4, V58.49, 404.11, 593.9, 592, 585, 759.1, 753.11, E879.1, 753.15, 585.9, 274, 588.8, 403.91, 404.9, 404.91, 404.92, 403, 404.01, 779.8, 250.4, 227, 227, 227, 404.93, 996.81, 593.89, 753.8, 593.2, 592, 753, 996.81, 223, 589.1, 996.81, 582.9, 585.9, 996.81, 753.1, 753.3, E878.0, 593.9, 753.3, 753.17, 583.9, 593.9, 589, 866 |
| Endocrine | 202.8, 200.1, 200.12, 201.9, 204, 202.88, 200.18, 196, 204.01, 785.6, 200.11, 200.13, 202.8, 785.6, 202.85, 202.81, 202.8, 202.8, 196, 196.9, 202.8, 202.82, 785.6, 202.8, 202, 202.87, V10.79, 785.6, 202.84, 202.01, 196.8, 457.1, 12.1, 785.6, V10.79, 196.5, 196.2, 238.7, 196.1, 200.14, 457.2, 238.7, V10.61, 201.9, 457, 202.8, 289.3, 245.2, 238.7, 785.6, 457.9, 785.6, 202.93, 196.9, 202.97, 757, V10.71, 288.8, 204.1, 202.83, 457.1, 289.3, 785.6, V77.9, 237.4, 239.7, 198.89, 623.5, 259.9, 200.2, V10.71 |
| Diabetes mellitus | 251.2, 362.01, 362.02, 250.4, 250.82, 790.2, 790.6, 250.5, 250.5, 250.6, 357.2, 790.2, 250, 250.4, 250.6, 250.8, 250.51, 250.5, 250.8, 250.82, 250.51, 250.51, V77.1, 250.12, 251.2, 250.12, 250.5, 250.83, 251.2, 250.41, 251.1, 250.52, 250.5, 648.81, 250.43, 250.53, 250.53, 250.81, 250.22, 250.13, 250.22, 250.83, 250.41, 250.5, 250.52, 250.52, 250.82, 253.5, V18.0, 588.1 |
| Hypertension | 401.9, 401.9, 250.82, 790.6, 401.9, 401.9, 250.82, 796.2, 402.9, 250.83, 405.99, 642.93, 642.01, 642.91, 401.9, E942.6, 405.09, 403.9, 437.2, 401, 401.1, 401, 642.33, 348.2, 779.8, 365.04, 572.3, 416, 416, 405.91, 416.8, 642.3 |
| Gastrointestinal | 153.3, 154.1, 153.9, 569.89, 154, 153.1, 578.9, 560.9, 569.3, 537.89, 558.9, 562.1, 153.6, 239, 532.3, 569.89, 532.7, 535.6, 558.9, 38.42, 569.89, 8.45, 153.2, 569.49, 79.89, 532.9, V58.11, 569, 154.1, 41.4, 537.89, 152.1, 578.9, V10.05, 787.8, 197.4, 535.5, V10.06, 9, 569.83, 569.6, 153.4, 560.9, 537.3, 41.04, 569.84, 239, 569.81, 8.8, 535, 560.9, 532, V45.89, V12.72, 532.4, V10.09, 560.81, 235.2, 38.49, 8.45, 235.2, 532.9, 569.81, 537.89, 557.9, 569.41, 997.4, 14.8, 787.99, 8.46, 535.5, 569.41, 997.4, 578.9, 569.82, 537.9, 560.1, 569.82, 557.9, 211.3, 556.9, 562, 558.9, 578.9, 536.9, 8.46, 535.6, 566, V71.9, 569.49, 564.3, V44.4, 569.89, 564.8, 8.46, 569.83, 997.4, 997.4, 997.4, 562.11, 211.2, 9.1, 211.3, 8.47, 8.5, 211.3, 569.83, 532.1, 535.61, 560, 569.83, 565.1, 619.1, 152.9, 568, 566, 569.43, 152, 8.46, 562.11, 8.61, 569.83, 569.83, 569.81, 596.1, 535.5, 151.4, 151.9, 151.5, 151.8, 151.1, 456.8, 531.7, 535.4, 531.3, V15.2, 537.89, 211.1, 531.9, V10.04, 235.2, 531, 531.4, 456.8, 535.1, 151.3, 230.2, 151.6, 535, 211.1, 536.3, 535, V10.04, 535.51, 578.9, 531.1, 535.1, 456.8, 531.5, 537.84, 535.01, 530.7, 535.1, 535.2, 535.5, 537.6, 202.83, 535.1, 531.4, 558.9, 558.9, 558, 569.85, 153, 555.1, 562.1, 562.13, 562.11, 562.12, 569.83, V76.49, 560.2, 230.4, 569.3, 154 |
| Stroke/TIA | 434.11, 434.91, 433.01, 433.11, 433.21, 433.31, 433.81, 433.91, 434.01, 434.91, 436, 435, 430, 431, 432, 432.1, 432.9, 853, 852, 852.01, 852.02, 852.03, 852.04, 852.05, 852.06, 852.07, 852.08, 852.09, 852.2, 852.21, 852.22, 852.23, 852.24, 852.25, 852.26, 852.27, 852.28, 852.29, 852.4, 852.41, 852.42, 852.43, 852.44, 852.45, 852.46, 852.47, 852.48, 852.49, I64, 992, 992, 434.01, 433, 331, 851, 434, 434.1, 348.5, 747.81, 414.9, 457.9, 720, 681.1, 473.9, 290.3, 524.02, 720, 414.9, 215.3 |
| IHD | 410.01, 410.02, 410.1, 410.11, 410.12, 410.2, 410.21, 410.22, 410.3, 410.31, 410.32, 410.4, 410.41, 410.42, 410.5, 410.51, 410.52, 410.6, 410.61, 410.62, 410.7, 410.71, 410.72, 410.8, 410.81, 410.82, 410.9, 410.91, 410.92, 411, 411.1, 411.8, 411.81, 411.89, 413, 413.1, 413.9, 414, 414.01, 414.02, 414.03, 414.04, 414.05, 414.06, 414.07, 414.1, 414.11, 414.12, 414.19, 414.2, 414.3, 414.4, 414.8, 414.9 |
| Gastrointestinal bleeding | 531, 531.2, 531.4, 531.6, 532, 532.2, 532.4, 532.6, 533, 533.2, 533.4, 533.6, 534, 534.2, 534.4, 534.6, 535.01, 535.11, 535.21, 535.31, 535.41, 535.51, 535.61, 535.71, 562.02, 562.03, 562.12, 562.13, 569.3, 569.85, 569.86, 578, 578.1, 578.9 |
| Heart failure | 57.9, 79.99, 198.5, 211.4, 225.4, 288.3, 303.02, 389.11, 398.9, 402.91, 416.9, 427.5, 428, 428.1, 428.9, 429.4, 478.31, 611.79, 617.3, 621.8, 711.09, 727.64, 784.4, 891.1, 916.9, V22.1, 428.99 |
| AMI | 386.11, 410, 410, 410, 410, 410.01, 410.01, 410.02, 410.02, 410.1, 410.1, 410.1, 410.11, 410.11, 410.12, 410.12, 410.2, 410.2, 410.2, 410.21, 410.21, 410.22, 410.22, 410.3, 410.3, 410.3, 410.31, 410.31, 410.32, 410.32, 410.4, 410.4, 410.4, 410.41, 410.41, 410.42, 410.42, 410.5, 410.5, 410.5, 410.51, 410.51, 410.52, 410.52, 410.6, 410.6, 410.6, 410.61, 410.61, 410.62, 410.62, 410.7, 410.7, 410.7, 410.71, 410.71, 410.72, 410.72, 410.8, 410.8, 410.8, 410.81, 410.81, 410.82, 410.82, 410.9, 410.9, 410.9, 410.91, 410.91, 410.92, 410.92, 411, 478.7, 501, 758, 830 |
| Hemorrhagic stroke | 431 |
| Pulmonary embolus and deep vein thrombosis | 453/415.1 |

**Supplementary Table 2. Baseline characteristics between patients with TIA/Istroke and hemorrhagic stroke before propensity score match**

* for SMD$\leq$0.2

| **Characteristics** | **Outcome TIA/Istroke (N=91) Mean(SD);Max;N or Count(%)** | **Outcome hemorrhagic stroke (N=117) Mean(SD);Max;N or Count(%)** | **SMD** |
| --- | --- | --- | --- |
| Male gender | 42(46.15%) | 71(60.68%) | 0.29 |
| Baseline age, year | 73.15(11.45);93.0;n=91 | 74.08(10.58);92.0;n=117 | 0.08* |
| Edoxaban | 10(10.98%) | 7(5.98%) | 0.18* |
| Edoxaban duration, days | 408.2(186.02);774.0;n=10 | 659.0(753.67);2289.0;n=7 | 0.46 |
| Warfarin duration, days | 354.22(151.41);1001.0;n=81 | 353.05(140.18);936.0;n=110 | 0.01* |
| Respiratory | 28(30.76%) | 49(41.88%) | 0.23 |
| Renal | 17(18.68%) | 34(29.05%) | 0.25 |
| Endocrine | 2(2.19%) | 1(0.85%) | 0.11* |
| Diabetes mellitus | 9(9.89%) | 22(18.80%) | 0.26 |
| Hypertension | 37(40.65%) | 48(41.02%) | 0.01* |
| Gastrointestinal | 18(19.78%) | 14(11.96%) | 0.22 |
| Congestive heart failure | 4(4.39%) | 3(2.56%) | 0.10* |
| Intracranial hemorrhage | 7(7.69%) | 45(38.46%) | 0.78 |
| PVD | 2(2.19%) | 4(3.41%) | 0.07* |
| AMI | 10(10.98%) | 19(16.23%) | 0.15* |
| SCD | 16(17.58%) | 28(23.93%) | 0.16* |
| HF | 41(45.05%) | 54(46.15%) | 0.02* |
| Baseline IHD | 17(18.68%) | 30(25.64%) | 0.17* |
| Baseline hemorrhagic stroke | 3(3.29%) | 0(0.00%) | 0.26 |
| Baseline gastrointestinal bleeding | 1(1.09%) | 2(1.70%) | 0.05* |
| Baseline TIA/Istroke | 0(0.00%) | 9(7.69%) | 0.41 |
| Charlson score | 3.53(1.59);11.0;n=91 | 3.85(1.91);11.0;n=117 | 0.19* |
| ACEI | 11(12.08%) | 14(11.96%) | 0.00* |
| ARB | 6(6.59%) | 7(5.98%) | 0.03* |
| Calcium channel blockers | 12(13.18%) | 31(26.49%) | 0.34 |
| Beta blockers | 14(15.38%) | 23(19.65%) | 0.11* |
| Diuretics for heart failure | 18(19.78%) | 24(20.51%) | 0.02* |
| Diuretics for hypertension | 0(0.00%) | 2(1.70%) | 0.19* |
| Nitrates | 5(5.49%) | 6(5.12%) | 0.02* |
| Antihypertensive drugs | 6(6.59%) | 12(10.25%) | 0.13* |
| Statins and fibrates | 12(13.18%) | 21(17.94%) | 0.13* |
| Antihyperlipidemic | 11(12.08%) | 21(17.94%) | 0.16* |
| Anticoagulants | 34(37.36%) | 55(47.00%) | 0.20* |
| K/Potassium, mmol/L | 4.06(0.49);5.1;n=57 | 4.07(0.52);5.9;n=80 | 0.01* |
| Urate, mmol/L | 0.45(0.18);0.92;n=27 | 0.41(0.12);0.71;n=31 | 0.23 |
| Albumin, g/L | 36.35(6.4);51.0;n=83 | 35.28(6.21);47.0;n=103 | 0.17* |
| Na/Sodium, mmol/L | 139.07(3.8);147.0;n=58 | 138.84(3.67);145.0;n=80 | 0.06* |
| Urea, mmol/L | 8.61(5.18);31.7;n=58 | 9.0(6.16);38.3;n=80 | 0.07* |
| Protein, g/L | 70.79(8.06);88.0;n=77 | 69.97(7.97);87.0;n=88 | 0.1* |
| Creatinine, umol/L | 134.52(129.57);853.0;n=58 | 162.34(218.16);1536.0;n=80 | 0.16* |
| Alkaline phosphatase, U/L | 89.52(55.21);378.0;n=83 | 96.49(56.02);428.8;n=103 | 0.13* |
| Aspartate transaminase, U/L | 23.01(8.25);40.0;n=21 | 39.82(68.3);411.0;n=33 | 0.35 |
| Alanine transaminase, U/L | 24.67(24.09);125.0;n=69 | 26.35(21.76);123.0;n=79 | 0.07* |
| Bilirubin, umol/L | 13.04(10.5);72.0;n=83 | 12.58(8.39);38.0;n=103 | 0.05* |
| HbA1c, g/dL | 12.82(1.69);14.9;n=10 | 12.31(1.98);15.1;n=7 | 0.27 |
| APTT, second | 32.81(8.27);59.6;n=45 | 33.84(9.37);81.6;n=69 | 0.12* |
| Prothrombin time, second | 16.09(7.7);47.5;n=36 | 17.55(9.1);58.9;n=57 | 0.17* |

**Supplementary Table 3. Baseline characteristics between patients with TIA/Istroke and hemorrhagic stroke after propensity score match (1:2)**

* for SMD$\leq$0.2

| **Characteristics** | **TIA/Istroke (N=58) Mean(SD);Max;N or Count(%)** | **Hemorrhagic stroke (N=55) Mean(SD);Max;N or Count(%)** | **SMD** |
| --- | --- | --- | --- |
| Male gender | 22(37.93%) | 34(61.81%) | 0.49 |
| Baseline age, year | 76.33(10.77);93.0;n=58 | 77.49(9.02);92.0;n=55 | 0.12* |
| Edoxaban | 2(3.44%) | 7(12.72%) | 0.35 |
| Edoxaban duration, days | 408.2(186.02);774.0;n=10 | 659.0(753.67);2289.0;n=7 | 0.46 |
| Warfarin duration, days | 330.94(143.85);1001.0;n=48 | 337.62(145.16);936.0;n=48 | 0.05* |
| Respiratory | 12(20.68%) | 21(38.18%) | 0.39 |
| Renal | 4(6.89%) | 15(27.27%) | 0.56 |
| Endocrine | 2(3.44%) | 0(0.00%) | 0.27 |
| Diabetes mellitus | 1(1.72%) | 4(7.27%) | 0.27 |
| Hypertension | 23(39.65%) | 23(41.81%) | 0.04* |
| Gastrointestinal | 9(15.51%) | 5(9.09%) | 0.20* |
| Congestive heart failure | 1(1.72%) | 4(7.27%) | 0.27 |
| Intracranial hemorrhage | 4(6.89%) | 14(25.45%) | 0.52 |
| PVD | 1(1.72%) | 1(1.81%) | 0.01* |
| AMI | 3(5.17%) | 4(7.27%) | 0.09* |
| SCD | 5(8.62%) | 8(14.54%) | 0.19* |
| HF | 22(37.93%) | 22(40.00%) | 0.04* |
| Baseline IHD | 5(8.62%) | 13(23.63%) | 0.42 |
| Baseline hemorrhagic stroke | 4(6.89%) | 0(0.00%) | 0.38 |
| Baseline gastrointestinal bleeding | 0(0.00%) | 0(0.00%) | - |
| Baseline TIA/Istroke | 0(0.00%) | 3(5.45%) | 0.34 |
| Charlson score | 3.67(1.19);6.0;n=58 | 3.98(1.41);7.0;n=55 | 0.24 |
| ACEI | 5(8.62%) | 2(3.63%) | 0.21 |
| ARB | 3(5.17%) | 5(9.09%) | 0.15* |
| Calcium channel blockers | 7(12.06%) | 10(18.18%) | 0.17* |
| Beta blockers | 9(15.51%) | 4(7.27%) | 0.26 |
| Diuretics for heart failure | 8(13.79%) | 10(18.18%) | 0.12* |
| Diuretics for hypertension | 0(0.00%) | 2(3.63%) | 0.27 |
| Nitrates | 4(6.89%) | 0(0.00%) | 0.38 |
| Antihypertensive drugs | 3(5.17%) | 3(5.45%) | 0.01* |
| Statins and fibrates | 9(15.51%) | 11(20.00%) | 0.12* |
| Antihyperlipidemic | 8(13.79%) | 11(20.00%) | 0.17* |
| Anticoagulants | 17(29.31%) | 18(32.72%) | 0.07* |
| K/Potassium, mmol/L | 4.02(0.35);4.6;n=37 | 4.06(0.44);4.8;n=36 | 0.11* |
| Urate, mmol/L | 0.45(0.2);0.92;n=15 | 0.42(0.15);0.71;n=17 | 0.15* |
| Albumin, g/L | 35.7(6.06);44.0;n=52 | 36.02(5.2);47.0;n=48 | 0.06* |
| Na/Sodium, mmol/L | 140.32(2.55);146.0;n=37 | 138.89(4.05);145.0;n=36 | 0.42 |
| Urea, mmol/L | 7.85(4.35);20.1;n=37 | 8.88(5.37);21.7;n=36 | 0.21 |
| Protein, g/L | 70.49(8.29);87.9;n=49 | 70.24(7.21);81.0;n=40 | 0.03* |
| Creatinine, umol/L | 106.43(55.7);285.0;n=37 | 126.58(73.98);328.0;n=36 | 0.31 |
| Alkaline phosphatase, U/L | 87.0(42.27);224.0;n=52 | 89.65(41.92);299.8;n=48 | 0.06* |
| Aspartate transaminase, U/L | 23.29(9.84);40.0;n=14 | 28.56(11.53);68.0;n=20 | 0.49 |
| Alanine transaminase, U/L | 27.38(31.08);125.0;n=44 | 28.58(28.86);123.0;n=37 | 0.04* |
| Bilirubin, umol/L | 12.07(7.16);33.4;n=52 | 13.49(9.74);35.4;n=48 | 0.17* |
| HbA1c, g/dL | 12.82(1.69);14.9;n=10 | 12.31(1.98);15.1;n=7 | 0.27 |
| APTT, second | 36.36(10.16);59.6;n=34 | 32.69(7.81);50.6;n=30 | 0.41 |
| Prothrombin time, second | 17.78(7.39);36.2;n=24 | 14.86(4.83);35.6;n=24 | 0.47 |
